# No Association between HPV Vaccination and Infertility in U.S. Females 18-33 Years Old

**DOI:** 10.1101/19012344

**Authors:** Katherine E. Mooney, Nicholas B. Schmuhl, Xiao Zhang, Laura G. Cooney, James H. Conway, Noelle K. LoConte

## Abstract

**Background:** Human papillomavirus (HPV) vaccines have been recommended as primary prevention of HPV-related cancers for over 10 years in the United States, and evidence reveals decreased incidence of HPV infections following vaccination. However, concerns have been raised that HPV vaccines could decrease fertility. This study examined the relationship between HPV immunization and self-reported infertility in a nationally representative sample.

**Methods:** Data from the 2013-2016 National Health and Nutrition Examination Survey were analyzed to asses likelihood of self-reported infertility among women aged 20 to 33, who were young enough to have been offered HPV vaccines and old enough to have been queried about infertility (n=1,114). Two logistic regression models, stratified by marital history, examined potential associations between HPV vaccination and infertility. Model 1 assessed the likelihood of infertility among women who had never been pregnant or whose pregnancies occurred prior to HPV vaccination. Model 2 accounted for the possibility of latent and/or non-permanent post-vaccine infertility by including all women 20-33 years old.

**Results:** 8.1% reported any infertility. Women who had ever been married and had received an HPV vaccine were **less** likely to report infertility (OR 0.04, 95% CI 0.01-0.57) in model 1. No other associations between HPV and infertility were found.

**Conclusion:** There was no evidence of increased infertility among women who received the HPV vaccine. These results provide further evidence of HPV vaccine safety and should give providers confidence in recommending HPV vaccination. Further research should explore protective effects of HPV vaccines on female and male fertility.

**What is already known on this subject?:** Despite evidence that HPV vaccines are safe and effective, concerns persist regarding a purported link between HPV vaccination and infertility. These concerns were refuted by a recent population-based cohort study that found no association between the HPV vaccine and primary ovarian insufficiency.

**What this study adds?:** This study broadens the existing evidence by exploring possible associations between HPV vaccination and any self-reported infertility. There was no evidence of infertility among women 20-33 years old who received one or more doses of HPV vaccine. This result provides further evidence of HPV vaccine safety, diminishing remaining concerns among clinicians and the public.

## INTRODUCTION

Human papillomavirus (HPV) is an extremely common sexually transmitted infection[1] and is linked to cervical, vulvar, vaginal, anal, and oropharyngeal cancers in females and oropharyngeal, anal, and penile cancers in males.[2] The HPV vaccine has been recommended for adolescents and young adults as primary prevention for HPV-related cancers for over 10 years,[3] and a recent Cochrane review of 26 randomized controlled trials (RCTs) found that the vaccine is both effective and safe.[4] While HPV vaccination rates are improving, they still lag behind other recommended adolescent vaccines.[5] HPV vaccine rates fall victim to some of the same barriers that adversely impact uptake of other vaccines – including access disparities, inadequate healthcare provider recommendations, and poor vaccine knowledge and attitudes.[6] Other barriers appear unique to the HPV vaccine, which prevents cancer-causing sexually transmitted infections. Notably, a number of studies have revealed parental[7,8] and physician[9,10] anxieties about associations between the HPV vaccine and sexual behavior, which are not supported by evidence.[11]

Recently, a small number of case reports[12,13] and anecdotes in the popular press[14,15] have given rise to concerns that the HPV vaccine causes lowered fertility by inducing primary ovarian insufficiency (POI). These concerns were refuted by a recent population-based cohort study of nearly 200,000 women that found no association between the HPV vaccine and POI.[16] However, POI is not the only condition implicated in female infertility. Thus, the purpose of this study is to explore potential associations between HPV vaccination and infertility. We selected the National Health and Nutrition Examination Survey (NHANES[17]) as the data source for this inquiry as it provides nationally representative data inclusive of individuals’ immunization and reproductive health histories. If females vaccinated against HPV report higher rates of infertility than those not vaccinated, more specific investigation of links between the HPV vaccine and conditions that cause infertility would be warranted.

## METHODS

NHANES has collected data to assess the health and nutritional status of adults and children in the U.S. intermittently since 1960, and continuously since 1999. The survey utilizes a representative sample of about 5,000 people living in various counties across the country. NHANES has long inquired about reproductive health, including pregnancy. In 2007-2008, NHANES first included questions about the HPV vaccine, and continues to do so. In 2013, the program began to include questions about difficulty or inability to become pregnant. In order to investigate potential associations between HPV vaccination and infertility, we analyzed data from two survey periods of the NHANES study: 2013-2014 (the first period that inquired about infertility) and 2015-2016 (the most recently available period). We confined our sample to women between 20 and 33 years old (N=1114) for two reasons: 1) NHANES did not query women younger than 20 if they had ever been pregnant; and 2) women older than 33 years old at the time of the 2013-2014 period would not have received the HPV vaccine, which was introduced in 2006. Because NHANES data are publicly available, this investigation was exempt from Institutional Review Board procedures.

### Outcome of interest: infertility

Difficulty or inability to become pregnant can be attributed to various factors, including age and disease. Infertility is defined as a failure to become pregnant after 12 months of regular and unprotected sexual intercourse.[18] Primary infertility occurs when a woman meets the definition of infertility and has never been pregnant. Secondary infertility occurs when a woman has been pregnant in the past, but is later unable to become pregnant.[19]

NHANES specifically asks female participants whether they are “pregnant **now**?” and whether they have “**ever** been pregnant?” with the specification that ever being pregnant includes a current pregnancy, live births, miscarriages, stillbirths, tubal pregnancies, and abortions. While a history of pregnancy suggests fertility, those who have never been pregnant are not necessarily infertile. Therefore, we used the following question, introduced in the 2013-2014 period of NHANES data collection, as a closer proxy for infertility:

> *[Has participant] ever attempted to become pregnant over a period of at least a year without becoming pregnant? (yes/no)*

The purpose of this study was to assess whether the HPV vaccine is a possible cause of infertility. While the HPV vaccine is recommended prior to sexual activity and during an age-range when most women or girls are not attempting to become pregnant, we account for both early pregnancy and late-adopters of the vaccine. In our first analysis, we assess the likelihood of reporting a 12-month period of infertility among women 20-33 years old who have never been pregnant (primary infertility), as well as women whose only pregnancies occurred prior to the age at which they received a first dose of the HPV vaccine (post-vaccine infertility).

Similarly, if the HPV vaccine were related to infertility, it could be hypothesized that the effect would be permanent, as would certainly be the case if the mechanism of infertility were POI; however, the effect might also be shorter-term. The effect of the vaccine on fertility could also be either immediate or delayed. To account for the possibility of latent and/or non-permanent, post-vaccine infertility, we performed a second analysis of all women 20-33 years old who reported any 12-month period of infertility, whether or not they had become pregnant at another time (i.e. primary or secondary infertility).

### Hypothesized predictor of infertility: any doses of HPV vaccine

As the main predictor variable, we used responses to the question,

> *[Has participant]* ***ever*** *received one or more doses of the HPV vaccine?*

This question has been asked of female NHANES participants between the ages of 9 and 59 since 2007.

### Other factors

During the two periods analyzed for this study, NHANES tracked several other health factors relevant to fertility, including body mass index (BMI), ever using birth control pills, and history of several sexually transmitted infections (STI). For the following analyses, we created a binary variable gauging whether an individual had any history of STI, based on self-report or laboratory results included in NHANES, as demonstrated by Anyalechi and colleagues.[20] Other covariates were health insurance status, routine access to healthcare, and socio-demographics, including age, race/ethnicity, marriage, education, and income. In addition to controlling for the aforementioned factors, we excluded women who reported having had their uterus or both ovaries removed, as both procedures result in a loss of fertility.

While recurrent miscarriage is a distinct condition from infertility, it can be related to some of the same underlying factors, and thus may be an important variable in the relationship between HPV vaccine and infertility. Unfortunately, NHANES does not ask explicitly about miscarriages; however, we attempted to account for miscarriage by controlling for history of live birth alongside history of pregnancy in our analysis of women who report any infertility. In multivariable models, any prior pregnancy and history of live birth were co-linear so only pregnancy history was retained in the final model.

### Statistical analysis

We calculated descriptive statistics (i.e. percentages, means, and standard deviations) for self-reported infertility, the receipt of HPV vaccination, and other factors. Survey weights provided by NHANES were used to account for the complex sampling design and response rates and to generate estimates at the population level. Multivariable logistic regression analyses were performed to examine the association between self-reported infertility and HPV vaccination status. Analyses were stratified by marital status based on the assumption that women who have never been married may be less likely to have attempted to become pregnant. All the regression models were adjusted for sociodemographic characteristics and other health- and healthcare-related factors. Due to multicollinearity between the history of prior pregnancy and live birth (see Table 1), the variable of the history of live birth was excluded from the regression models. All analyses were conducted with the software STATA/SE 14.2 (StataCorp LP, College Station, TX).

**Table 1.** Descriptive statistics of females aged 20-33: National Health and Nutrition Examination Survey, 2013-2016 (N=1114)

|  | No self-reported infertility (N=1018) | Self-reported infertility (N=96) | P-value* |
| --- | --- | --- | --- |
| Overall, % | 91.9 | 8.1 |  |
| **Personal characteristics** |  |  |  |
| Age, mean (SD) | 25.3 (4.4) | 27.8 (4.0) | **<0.001** |
| Race, % |  |  | 0.284 |
| NH white | 57.8 | 56.0 |  |
| NH black | 13.1 | 16.7 |  |
| Hispanic | 17.9 | 20.9 |  |
| Other | 11.2 | 6.4 |  |
| College education, % | 32.6 | 21.8 | 0.079 |
| Ever married, % | 55.1 | 78.3 | **<0.001** |
| Ratio of family income to poverty, % | 2.5 (1.6) | 2.1 (1.4) | **0.034** |
| **Health related factors** |  |  |  |
| Receipt of HPV vaccination, % | 36.1 | 20.4 | **0.01** |
| BMI category, % |  |  | **<0.001** |
| Underweight or normal | 41.9<br>25.8 | 28.6<br>14.0 |  |
| Overweight | 32.3 | 57.4 |  |
| Obese |  |  |  |
| Had ever taken birth control pills, % | 70.3 | 63.2 | 0.196 |
| Had any sexually transmitted infections, % | 18.2 | 17.3 | 0.840 |
| History of pregnancy, % | 50.9 | 78.7 | **<0.001** |
| History of live birth, % | 50.8 | 78.7 | **<0.001** |
| **Healthcare related factors** |  |  |  |
| Covered by health insurance, % | 81.0 | 65.1 | **0.001** |
| Had a routine place to go for healthcare, % | 78.4 | 82.3 | 0.422 |

\*P-values are based on t-test for continuous variables and chi-square test for categorical variables.

## RESULTS

Overall, 8.1% of our sample reported having ever experienced a 12-month period of infertility. The rate of primary infertility (i.e. among women who had never been pregnant) is 3.7%, and the rate of secondary infertility (i.e. among women who had been pregnant in the past) is 12.0%. In general, females who reported infertility during the past 12 months differed from their counterparts in several ways. Those with a history of infertility were slightly older (27.8 years vs. 25.3 years; p<0.001), more likely to be obese (57.4% vs. 32.1%; p <0.001) and more likely to have ever been married (p<0.001). They also had a lower income-to-poverty ratio (2.1 vs. 2.6; p=0.019) and were less likely to have health insurance (65.1% vs. 81.0%, p=0.001). Women who reported infertility had a higher rate of history of past pregnancy (78.7% vs. 50.9%, p<0.001) and livebirth (78.7% vs. 50.8%, p<0.001), but were less likely to have received at least one dose of the HPV vaccine (20.4% vs. 36.4%; p=0.02). See table 1.

### Primary and Post-vaccine infertility

Our first analysis included women with primary or post-vaccine infertility defined as women who had never been pregnant, or who had not been pregnant since receiving the HPV vaccine and reported a year of attempting pregnancy without success. Multivariable models indicate that there is no association between HPV vaccination and infertility among women in this population who have never been married (OR 0.91, 95% CI 0.24-3.49), or for the overall sample (OR 0.48, 95% CI 0.17-1.35). Among women who had ever been married, those who had been vaccinated against HPV were less likely to report infertility (OR 0.04, 95% CI 0.01-0.57). BMI was the most significant predictor of infertility in our model, with obese women nearly five times more likely to report infertility (OR 4.72, 95% CI 1.41-15.8) compared to women with underweight or normal BMI. Race also played a role, as Black women were four times more likely to report infertility (OR 4.01, 95% CI 1.05-15.3) than White women. Women with a college education (OR 0.11, 95% CI 0.01-0.97) and women covered by health insurance (OR 0.23, 95% CI 0.08-0.67) were less likely than their counterparts to report infertility, though health insurance coverage was not a significant predictor among never married women. Age, income/poverty ratio, ever use of birth control pills, history of STI, and routine access to health care were not individually associated with the likelihood of reporting infertility. See table 2.

**Table 2.** Predictors of primary and post-vaccine infertility^1^ among females aged 20-33, National Health and Nutrition Examination Survey, 2013-2016

|  | Overall<br>(N=505) |  | Ever married<br>(N=184) |  | Never married<br>(N=321) |  |
| --- | --- | --- | --- | --- | --- | --- |
|  | OR | 95% CI | OR | 95% CI | OR | 95% CI |
| HPV vaccination | 0.48 | 0.17-1.35 | **0.04** | **0.01-0.57** | 0.91 | 0.24-3.49 |
| Age | 1.11 | 0.98-1.27 | 0.98 | 0.76-1.27 | 1.05 | 0.88-1.24 |
| Race |  |  |  |  |  |  |
| NH white | Ref |  | Ref |  | Ref |  |
| NH black | **4.01** | **1.05-15.3** | --* | -- | 4.90 | 0.86-27.9 |
| Hispanic | 1.10 | 0.30-4.10 | 2.52 | 0.39-16.2 | 0.48 | 0.04-6.54 |
| Other | -- | -- | --* | -- | --* | -- |
| College | **0.11** | **0.01-0.97** | 0.48 | 0.03-7.13 | --* | -- |
| education |  |  |  |  |  |  |
| Ever married | 2.51 | 0.86-7.35 | NA |  | NA |  |
| Ratio of family income to poverty | 0.78 | 0.52-1.19 | 0.84 | 0.39-1.82 | 0.71 | 0.40-1.26 |
| BMI category |  |  |  |  |  |  |
| Underweight or normal | Ref |  | --* | -- | Ref |  |
| Overweight | 0.96 | 0.19-4.84 |  |  | 0.72 | 0.11-4.70 |
| Obese | **4.72** | **1.41-15.8** |  |  | 1.97 | 0.45-8.63 |
| Had ever taken birth control pills | 0.80 | 0.30-2.15 | 0.31 | 0.04-2.29 | 0.64 | 0.16-2.59 |
| Had any sexually transmitted infections | 1.72 | 0.44-6.64 | --* | -- | 3.69 | 0.74-18.5 |
| Covered by health insurance | **0.23** | **0.08-0.67** | **0.06** | **0.01-0.44** | 0.55 | 0.12-2.53 |
| Had a routine place to go for healthcare | 0.82 | 0.28-2.39 | 0.70 | 0.07-6.70 | 0.93 | 0.21-4.23 |

1 Primary infertility refers to infertility reported by women who have never been pregnant, and post-vaccine infertility refers to infertility reported by those whose only pregnancies occurred prior to the age at which they received a first dose of the HPV vaccine.
\*Variables were dropped due to perfect prediction.
Bold values represent statistical significance at the 0.05 level.

### Any infertility

When including all women between the ages of 20 and 33 in these two periods of the NHANES sample, we did not find a significant relationship between HPV vaccination and infertility. In this sample, women who had ever been married were more than twice as likely to report any 12-month period of infertility as those who had not been married (OR 2.23, 95% CI 1.28-3.89). After stratification by marital status, we found that among never married women, those with higher incomes were less likely to report infertility (OR 0.65, 95% CI 0.45-0.95). No other variables were associated with infertility in this subgroup. Married women with BMIs categorized as obese were more than twice as likely to have experienced infertility (OR 2.38, 95% CI 1.22-4.65), a pattern which persisted in the overall sample (OR 2.06, 95% CI 1.20-3.53). Among married women, those covered by health insurance were less likely to report infertility (OR 0.34, 95% CI 0.18-0.66), and this pattern persisted in the overall sample (OR 0.50, 95% CI 0.30-0.84). See table 3.

**Table 3.** Predictors of any infertility among females aged 20-33, National Health and Nutrition Examination Survey, 2013-2016

|  | Overall<br>(N=1114) |  | Ever married<br>(N=608) |  | Never married<br>(N=506) |  |
| --- | --- | --- | --- | --- | --- | --- |
|  | OR | 95% CI | OR | 95% CI | OR | 95% CI |
| HPV vaccination | 0.67 | 0.38-1.20 | 0.54 | 0.25-1.20 | 0.85 | 0.34-2.09 |
| Age | 1.03 | 0.97-1.10 | 1.04 | 0.95-1.12 | 0.99 | 0.88-1.11 |
| Race |  |  |  |  |  |  |
| NH white | Ref |  | Ref |  | Ref |  |
| NH black | 1.31 | 0.70-2.44 | 0.92 | 0.37-2.26 | 1.70 | 0.61-4.73 |
| Hispanic | 0.76 | 0.42-1.40 | 0.85 | 0.42-1.71 | 0.50 | 0.12-2.19 |
| Other | 0.55 | 0.24-1.27 | 0.62 | 0.25-1.53 | --* | -- |
| College education | 0.78 | 0.40-1.53 | 1.13 | 0.52-2.47 | --* | -- |
| Ever married | **2.23** | **1.28-3.89** | NA |  | NA |  |
| Ratio of family income to poverty | 0.94 | 0.79-1.13 | 1.06 | 0.85-1.32 | **0.65** | **0.45-0.95** |
| BMI category |  |  |  |  |  |  |
| Underweight or normal | Ref |  | Ref |  | Ref |  |
| Overweight | 0.69 | 0.34-1.39 | 0.53 | 0.23-1.30 | 1.11 | 0.34-3.62 |
| Obese | **2.06** | **1.20-3.53** | **2.38** | **1.22-4.65** | 1.86 | 0.71-4.90 |
| Had ever taken birth control pills | 0.87 | 0.53-1.41 | 0.81 | 0.44-1.49 | 0.97 | 0.39-2.36 |
| Had any sexually transmitted infections | 1.26 | 0.68-2.33 | 0.99 | 0.41-2.35 | 1.88 | 0.73-4.87 |
| History of pregnancy | 1.28 | 0.72-2.26 | 1.43 | 0.70-2.93 | 1.07 | 0.40-2.90 |
| Covered by health insurance | **0.50** | **0.30-0.84** | **0.34** | **0.18-0.66** | 1.08 | 0.41-2.85 |
| Had a routine place to go for healthcare | 1.59 | 0.86-2.93 | 1.89 | 0.89-4.05 | 0.98 | 0.33-2.88 |

\*Variables were dropped due to perfect prediction.
Bold values represent statistical significance at the 0.05 level.

## DISCUSSION

In both analyses, the first focusing on women with possible primary or post-vaccine infertility and the second attempting to take into account latent and/or non-permanent post-vaccine infertility and controlling for history of pregnancy, there was a consistent inverse relationship between HPV vaccination and self-reported infertility, although this association was only statistically significant among ever married women in analysis 1. Importantly, the analyses were conducted with a proxy for infertility that is nearly identical to how infertility is defined in clinical practice and controlled for important covariates like ever using birth control. Obtaining null results using an outcome variable that broadly defines infertility dispels concerns about association between HPV vaccination and infertility due to POI or any other condition. This lack of association may help allay public concerns about vaccine safety and bolster HPV vaccination rates. It should also help providers feel confident in making strong recommendations for HPV vaccination for adolescent girls and boys.

The statistically significant relationships between infertility and other variables in our analyses were largely expected. For example, the relationship between obesity and infertility, which was seen in both analyses, is well supported by prior data.[21] It should also be no surprise that women who have ever been married were more likely to report infertility (Table 3) as this population is presumably more likely to be trying to have children. Similarly, women with insurance coverage were less likely to report infertility than those without insurance in both samples analyzed (Tables 2 and 3), likely because this group had better access to medical care. Interestingly, increased odds of reporting infertility among Black women was seen in the first analysis (women who had never been pregnant, post-HPV vaccination), but not the larger second analysis of women who had ever experienced a 12-month period of fertility. Previous research has observed, but not explained, increased odds of infertility among Black women when controlling for common risk factors.[22]

There are several limitations that should be considered when interpreting the results of this study. First, the sample size was limited because survey questions about infertility and HPV immunization only coincided for two periods of the NHANES study (2013-2014 and 2015-2016). Similarly, the upper age limit for inclusion in this study was 33 years, because women older than 33 during the 2013-2014 NHANES survey would not have been eligible for the HPV vaccine when it was introduced in 2006.If possible, future research should include older women to explore the possibility of very delayed impacts of the HPV vaccine on fertility.

Second, all variables were self-reported, including HPV vaccination status and inability to become pregnant when desired. It is possible that questions of vaccination status in particular could be subject to recall bias since the many vaccinations given throughout childhood and adolescents are easily confused. However, previous research using self-reported receipt of =/> 1 dose of HPV vaccine (NHANES question IMQ060) have compared respondents’ answers to medical records and have documented 86%-87% sensitivity, 83%-87% specificity, and 70%-73% agreement.[23,24]

Among women without a history of infertility, the NHANES data did not allow us to distinguish those who had been successful in their attempts to become pregnant from those who had never tried to conceive. Another limitation was the inability to easily control for miscarriage, an important covariate that could be associated with infertility. Because miscarriage was not explicitly measured by NHANES, we were limited to a proxy constructed by comparing self-reported pregnancies and live births.

While only one subgroup in these analyses showed a significantly lower rate of infertility among those who had been vaccinated against HPV, ongoing research seeks to understand whether HPV vaccines are actually protective of fertility. A recent systematic review of more than 100 peer-reviewed articles published between 1994 and 2014 found that HPV *infections* were related to decreased reproductive function in both males and females. The studies reviewed focused largely on the association of HPV infection with semen parameters, failed *in vitro* fertilization, and poor pregnancy outcomes (e.g., miscarriage).[25] Given that the 9-valent HPV vaccine is 96% effective at preventing HPV infections and related conditions,[3] it is reasonable to believe that those who are vaccinated will have fewer fertility problems. Moreover, cancers and pre-cancers caused by HPV infection also restrict fertility. For example, cervical cancer workup and treatment (e.g. cervical conization) can result in cervical insufficiency, miscarriages, and preterm birth. Further research is needed to quantify the benefits of HPV vaccination to both female and male fertility. In the meantime, healthcare providers and patients can be reassured that there is no association between HPV vaccination and infertility.

## Data Availability

The authors confirm that the data supporting the findings of this study are available within the article and its supplementary materials.

## Acknowledgments

Funding (NL, XZ) received from the University of Wisconsin Carbone Cancer Center Support Grant NCI P30 CA014520

## Conflicts of interest

None

## Notes

### Competing Interest Statement

The authors have declared no competing interest.

### Funding Statement

Funding received from the University of Wisconsin Carbone Cancer Center Support Grant NCI P30 CA014520

## REFERENCES

1 Serrano B, Brotons M, Bosch FX, et al. Epidemiology and burden of HPV-related disease. Best Pract Res Clin Obstet Gynaecol 2018;47:14–26. doi:10.1016/j.bpobgyn.2017.08.006

2 Saslow D, Andrews KS, Manassaram-Baptiste D, et al. Human papillomavirus vaccination guideline update: American Cancer Society guideline endorsement. CA Cancer J Clin 2016;66:375–85. doi:10.3322/caac.21355

3 Petrosky E, Bocchini JA, Hariri S, et al. Use of 9-Valent Human Papillomavirus (HPV) Vaccine: Updated HPV Vaccination Recommendations of the Advisory Committee on Immunization Practices. MMWR Morb Mortal Wkly Rep 2015;64:300–4.

4 Arbyn M, Xu L, Simoens C, et al. Prophylactic vaccination against human papillomaviruses to prevent cervical cancer and its precursors. Cochrane Database Syst Rev Published Online First: 2018. doi:10.1002/14651858.CD009069.pub3

5 Walker TY, Elam-Evans LD, Yankey D, et al. National, Regional, State, and Selected Local Area Vaccination Coverage Among Adolescents Aged 13–17 Years — United States, 2018. Morb Mortal Wkly Rep 2019;68:718–23. doi:10.15585/mmwr.mm6833a2

6 Kessels SJM, Marshall HS, Watson M, et al. Factors associated with HPV vaccine uptake in teenage girls: A systematic review. Vaccine 2012;30:3546–56. doi:10.1016/j.vaccine.2012.03.063

7 Brewer NT, Fazekas KI. Predictors of HPV vaccine acceptability: A theory-informed, systematic review. Prev Med 2007;45:107–14. doi:10.1016/j.ypmed.2007.05.013

8 Gidengil C, Chen C, Parker AM, et al. Beliefs around childhood vaccines in the United States: A systematic review. Vaccine 2019;37:6793–802. doi:10.1016/j.vaccine.2019.08.068

9 Daley MF, Crane LA, Markowitz LE, et al. Human Papillomavirus Vaccination Practices: A Survey of US Physicians 18 Months After Licensure. PEDIATRICS 2010;126:425–33. doi:10.1542/peds.2009-3500

10 Kempe A, O’Leary ST, Markowitz LE, et al. HPV Vaccine Delivery Practices by Primary Care Physicians. Pediatrics 2019;144:e20191475. doi:10.1542/peds.2019-1475

11 Liddon NC, Leichliter JS, Markowitz LE. Human Papillomavirus Vaccine and Sexual Behavior Among Adolescent and Young Women. Am J Prev Med 2012;42:44–52. doi:10.1016/j.amepre.2011.09.024

12 Colafrancesco S, Perricone C, Tomljenovic L, et al. Human papilloma virus vaccine and primary ovarian failure: another facet of the autoimmune/inflammatory syndrome induced by adjuvants. Am J Reprod Immunol N Y N 1989 2013;70:309–16. doi:10.1111/aji.12151

13 Little DT, Ward HRG. Adolescent Premature Ovarian Insufficiency Following Human Papillomavirus Vaccination: A Case Series Seen in General Practice. J Investig Med High Impact Case Rep 2014;2:2324709614556129. doi:10.1177/2324709614556129

14 14 A note from the publisher | The Star. https://www.thestar.com/news/2015/02/20/a-note-from-the-publisher.html (accessed 28 Aug 2019).

15 Wahlberg D. Judge says HPV vaccine didn’t cause ovary failure in Mount Horeb sisters. madison.com. http://host.madison.com/news/local/health-med-fit/judge-says-hpv-vaccine-didn-t-cause-ovary-failure-in/article_5f5678fd-9fcd-5be1-9be6-0f33a0e6a211.html (accessed 13 Dec 2017).

16 Naleway AL, Mittendorf KF, Irving SA, et al. Primary Ovarian Insufficiency and Adolescent Vaccination. Pediatrics 2018;142:e20180943. doi:10.1542/peds.2018-0943

17 National Health and Nutrition Examination Survey. 2019. https://www.cdc.gov/nchs/nhanes/index.htm (accessed 30 Aug 2019).

18 Practice Committee of the American Society for Reproductive Medicine. Definitions of infertility and recurrent pregnancy loss: a committee opinion. Fertil Steril 2013;99:63. doi:10.1016/j.fertnstert.2012.09.023

19 Vander Borght M, Wyns C. Fertility and infertility: Definition and epidemiology. Clin Biochem 2018;62:2–10. doi:10.1016/j.clinbiochem.2018.03.012

20 Anyalechi GE, Hong J, Kreisel K, et al. Self-Reported Infertility and Associated Pelvic Inflammatory Disease Among Women of Reproductive Age—National Health and Nutrition Examination Survey, United States, 2013–2016. Sex Transm Dis 2019;46:446. doi:10.1097/OLQ.0000000000000996

21 Obesity and reproduction: a committee opinion. Fertil Steril 2015;104:1116–26. doi:10.1016/j.fertnstert.2015.08.018

22 Wellons MF, Lewis CE, Schwartz SM, et al. Racial differences in self-reported infertility and risk factors for infertility in a cohort of black and white women: The CARDIA Women’s Study. Fertil Steril 2008;90:1640–8. doi:10.1016/j.fertnstert.2007.09.056

23 Adjei Boakye E, Tobo BB, Osazuwa-Peters N, et al. A Comparison of Parent- and Provider-Reported Human Papillomavirus Vaccination of Adolescents. Am J Prev Med 2017;52:742–52. doi:10.1016/j.amepre.2016.10.016

24 Lewis RM, Markowitz LE. Human papillomavirus vaccination coverage among females and males, National Health and Nutrition Examination Survey, United States, 2007–2016. Vaccine 2018;36:2567–73. doi:10.1016/j.vaccine.2018.03.083

25 Souho T, Benlemlih M, Bennani B. Human Papillomavirus Infection and Fertility Alteration: A Systematic Review. PLOS ONE 2015;10:e0126936. doi:10.1371/journal.pone.0126936

